# Time-varying reproduction number estimation: Fusing compartmental models with generalised additive models

**DOI:** 10.1101/2024.03.26.24304928

**Authors:** Xiaoxi Pang, Yang Han, Elise Tessier, Nurin Abdul Aziz, Lorenzo Pellis, Thomas House, Ian Hall

**Affiliations:** University of Manchester; University of Nottingham; The University of Manchester; UKHSA

## Abstract

The reproduction number, the mean number of secondary cases infected by each primary case, is a central metric in infectious disease epidemiology, and played a key role in the COVID-19 pandemic response. This is because it gives an indication of the effort required to control the disease. Beyond the well-known *basic* reproduction number, there are two natural versions, namely the *control* and *effective* reproduction numbers. As behaviour, population immunity and viral characteristics can change with time, these reproduction numbers can vary over time and in different regions.

Real world data can be complex, for example with daily variation in numbers due to weekend surveillance biases as well as natural stochastic noise. As such, in this work we consider a Generalised Additive Model to smooth real data through the explicit incorporation of day-of-the-week effects, to provide a simple measure of the time-varying growth rate associated with the data.

Converting the resulting spline into an estimator for both the control and effective reproduction numbers requires assumptions on a model structure, which we here assume to be a compartmental model. The reproduction numbers calculated are based on both simulated and real world data, and are compared with estimates from an already existing tool.

The derived method for estimating the time-varying reproduction number is effective, efficient and comparable to other methods. It provides a useful alternative approach, which can be included as part of a toolbox of models, that is particularly apt at smoothing out day-of-the-week effects in surveillance.

## 1 Background and introduction

Severe Acute Respiratory Syndrome Coronavirus 2 (SARS-CoV-2) is the etiological agent of the Coronavirus Disease 2019 (COVID-19). The infection was first identified in China in late 2019, and only a few months later cases were found all around the world and a global pandemic was declared [1]. Due to the high transmission, rapid mutation and changing human behaviour, many countries and regions have experienced multiple outbreaks: for instance, the United Kingdom (UK) experienced a second wave in the winter of 2020 driven by Alpha variant; the first case of the Delta variant in the UK was found in mid-April of 2021 [2] and generated a new wave that summer; and in December 2021 the Omicron wave began [3]. In such situations, a method for estimation of the transmission rate over time is needed because time-varying factors influencing transmission will impact the effectiveness of the mitigation deployed. Therefore, such estimates can inform if stronger or weaker interventions may be required.

There are two commonly used indicators to assess the transmission impact of an epidemic, namely growth rate and reproduction number. The former describes the rate of change of the observed cases, which provides a statistical measure indicating if the infectious disease risk is increasing (positive) or decreasing (negative) [4, 5]. A method using Bayesian modelling with Gaussian processes to estimate the growth rate of SARS-CoV-2 in England has been developed [6]. Meanwhile, the reproduction number can be a more useful metric for decision makers as it can inform the extent of public interventions against the disease. For instance, if a proposed intervention was to reduce the infection rate by a factor equalling the reciprocal of reproduction number then this may be expected to be sufficient for an effective mitigation. Hence, obtaining the real-time reproduction number is also helpful to flexibly adjust the intervention policy [4].

The basic reproduction number is defined as the mean number of secondary cases infected by a primary case in an otherwise wholly susceptible population. For the simple compartmental SIR model it is defined as *R*_0_ = *β/γ*, where *β* is the infection rate (a composite of the average number of contacts and the probability of infection given a contact) and *γ* is the removal rate [7]. A systematic review [8] has done a full-text assessment of 23 studies estimating the basic reproduction number of SARS-CoV-2 early in the pandemic, obtaining an overall mean of 3.38 *±* 1.40, which is consistent with the challenges in controlling its rapid spread [5]. The fact that such a range of different estimates exist is not surprising: *R*_0_ can virtually never be measured directly, but rather needs to be estimated through the use of a model, and different assumptions on model structure and parameter values might lead to different estimates; *R*_0_ is not only a property of the virus, but also of the population characteristics and mixing patterns, and so it might naturally vary between different settings, geographical regions and populations; and finally, as it became evident during the COVID-19 pandemic, the intrinsic transmissibility of the virus is also not immutable, as variants might evolve which are more efficient at spreading and can outcompete the previous ones. Even when restricting to a setting and time where it is reasonable to assume a single value of *R*_0_, individual and societal behavioral responses may change when faced with a disease [9]. Following Pellis *et al* [4], the *effective* reproduction number *R*_*E*_(*t*) (often denoted by *R*_*t*_) describes the expected number of secondary infections under the current conditions of population mixing, transmission and immunity. The *control* reproduction number (or reproduction number excluding immunity), *R*_*C*_(*t*), describes the expected number of secondary infections under the current contact and transmission patterns in an otherwise fully susceptible population [4]. The control and effective reproduction numbers can vary with time and are context-specific so driven by the data used in the inference.

Previous work has considered methods to estimate time varying reproduction numbers. For example, before the COVID-19 pandemic studies modelled the reproductive rate by multiplicative random walk and used in stochastic SIR model to explore the Ebola outbreak [10]. Cori *et al* [11] give an approach to inferring the effective reproduction number over time from case data and information of serial interval (time difference of symptom onset between the infector and the infected individual), and the corresponding R package `EpiEstim` [12] has made inference of such a time-varying reproduction number accessible to wider user community. Subsequent studies work out the distribution of time-varying reproduction number conditional on serial interval, and surmises the reasons why the reproduction number is time-dependent, including but not necessarily disease control interventions [13], and `EpiEstim` has been updated to account for imported cases in the reproduction number estimation. Here we use *R* estimates from `EpiEstim` as a comparator for the method developed in this study. However, measuring the serial interval is not always straightforward, as evidenced for example by the variability observable in a systematic review [14].

Whilst this work had begun in 2020 (arising from ideas posited in [5]), we conducted a scoping literature search in December 2023 on PUBMED for recent similar articles. Searching (“Time varying” AND “Reproduction Number”) yielded 236 returns. Exclusion on title, abstract and full text (not respiratory disease, no explicit mention of time varying infection rate) gave 38 studies explicitly considering time varying reproduction number estimates. After a full read, we concluded that the majority of studies (31) had been cited above, directly used `EpiEstim` (or a variant of this method) or were applying alternative analyses based on renewal equations [15]. The developers of `EpiEstim` have conducted a literature review of recent applications [16]. In general these studies hinge on direct measurement of either the generation time or serial interval.

Another issue for modelling the SARS-CoV-2 outbreak is parametrisation of the effectiveness of pandemic interventions, because natural and mandated human behaviour change can influence disease transmission, so improper assumptions for behaviour change in the epidemic modelling will yield biases [17]. Modelling with a (semi-parametric) smoothing process can be an alternative option when mechanistic assumptions are unclear or unknown. Two studies from the literature search [18, 19] fuse deep learning techniques and Kalman filters, respectively, to compartmental models.

A generalised additive model (GAM) [20, 21] is an established statistical tool that can be applied on case data. In this study we propose a novel technique for inferring the reproduction number using GAMs to derive growth rates and, with structural assumptions on the mechanisms of transmission, to translate them to reproduction number estimates. The remaining 5 studies from the literature review have performed similar analyses. To infer a time-varying reproduction number Hong and Li apply cubic B-splines with Poisson Noise [22], Gressani *et al* extend this with a Negative Binomial noise model [23], whilst Eales *et al* use Bayesian P-splines and verify the result by comparing it with a simple exponential epidemic model [24]. Wood and Wit use adaptive smoothing splines [25] with Negative Binomial noise with a highly structured model. Gleeson *et al* employs a negative binomial noise term with a GAM using thin plate splines integrated with a compartmental model [26]. This study in similar in concept to ours, though our compartmental model has a different structure, as we are applying method to two different datasets from care home surveillance, and using the `mgcv` package [27] we can switch flexibly between splines (details below). Our approach also permits the use of day-of-the-week terms in regression to account for structured surveillance bias.

## 2 Materials and methods

Generalised additive models (GAMs) [28] have been used for rapid estimation of the instantaneous growth rates in support of policy decisions in the United Kingdom [29, 5]. However, reproduction number estimates can offer an insight on the amount of transmission that needs to be prevented to control spread, thus offering additional context to the calculated growth rate, but requires additional structural assumptions to be made. In this study new estimators of time-varying reproduction numbers are developed by combining GAMs (with time as the argument of the smoothing function) and compartmental transmission models based on ordinary differential equations.

### 2.1 Modelling the daily incidence time series

The daily case incidence data *C*_*t*_ is fitted by application of a generalised additive model (GAM) with log link assuming Negative Binomial error structure,

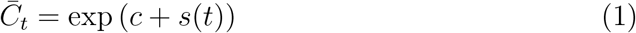

where *t* denotes time (throughout, we use day as the time unit), 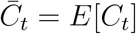 denotes the mean at time *t* while *c* and *s*(*t*) are the ‘intercept’ (constant over time) and the smooth function (spline) over time, respectively [20]. The expected value of the spline over time is zero (*E*[*s*(*t*)] = 0 over the length of the time series) and so 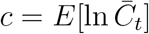 . For a generic function *f*, we use the *f*_*t*_ subscript notation to denote surveillance data on day *t* and *f* (*t*) to denote the continuous function we want to fit to such data.

Splines have been commonly used to apply a smoothing process to data. Here we use penalised splines, which involve the use of a roughness penalty, say 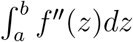, instead of the number of basis functions, to control the smoothness of the fitted curve [30]. However, other spline options available in the `mgcv` package (e.g. cubic spline and Gaussian Process) have been tested without dramatic influence on results (not shown).

Beyond the choice of which splines to use, the choice of knots has important repercussion on the resulting fitted function and problems may arise if they are too many or too few. In this paper we will use a spline with *k* = floor(*K/*20) knots where *K* is the total number of data points being fitted. The number of knots is chosen so that there is a knot for 20 days of data, which roughly corresponds to 3-4 generations of infection in the case of SARS-CoV-2. The rationale for this is that shorter gaps between knots may over-fit data and interact with the day of week terms whilst longer gaps between knots would over-smooth the data and inhibit detection of changes in trend due to variants and interventions.

Fitting a continuous function *C*(*t*) to discrete data *C*_*t*_ allows us to define an instantaneous growth rate at any time point *t*, usually denoted by *r*(*t*), which can be computed as the per-capita variation in the number of cases (i.e. *Ċ* (*t*)*/C*(*t*)). In terms of the elements of the GAM (see also [5]), this is given by

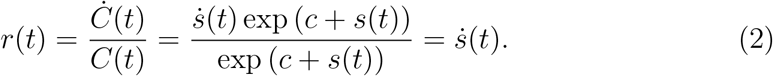

The intercept *c* may contain non-temporal fixed effects or random effects. Regular spikes in surveillance data (periodic every seven days) (Figures 5, top panel, and 6, top panel) are likely due to operational constraints in data reporting and collecting pattern or other systematic lags. Hence the generalised additive model, when fitted to care home data sets, should include a parametric component to account for day-of-the-week effects:

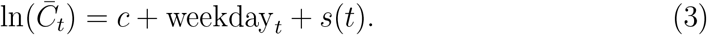

The estimation process will produce the mean and standard deviation given the data to generate confidence intervals. However, here a Bayesian credible interval (CrI) for the GAM predictions is used, following [28]. Initially the coefficients are simulated from the asymptotic multi-normal distribution, and this is multiplied with the design matrix. Though the asymptotic multi-normal distribution’s mean and covariance matrix can be simply extracted from model output, it is actually conditional on a fixed smoothing parameter, which has been determined in the model fitting. To investigate this issue, a Bayesian parametric simulation approach is used to generate unconditional credible interval [28]. In simulation and testing, the conditional and unconditional CrIs were not very different but the latter is more computationally expensive, so the results presented are based on the conditional method.

**Table 1:**
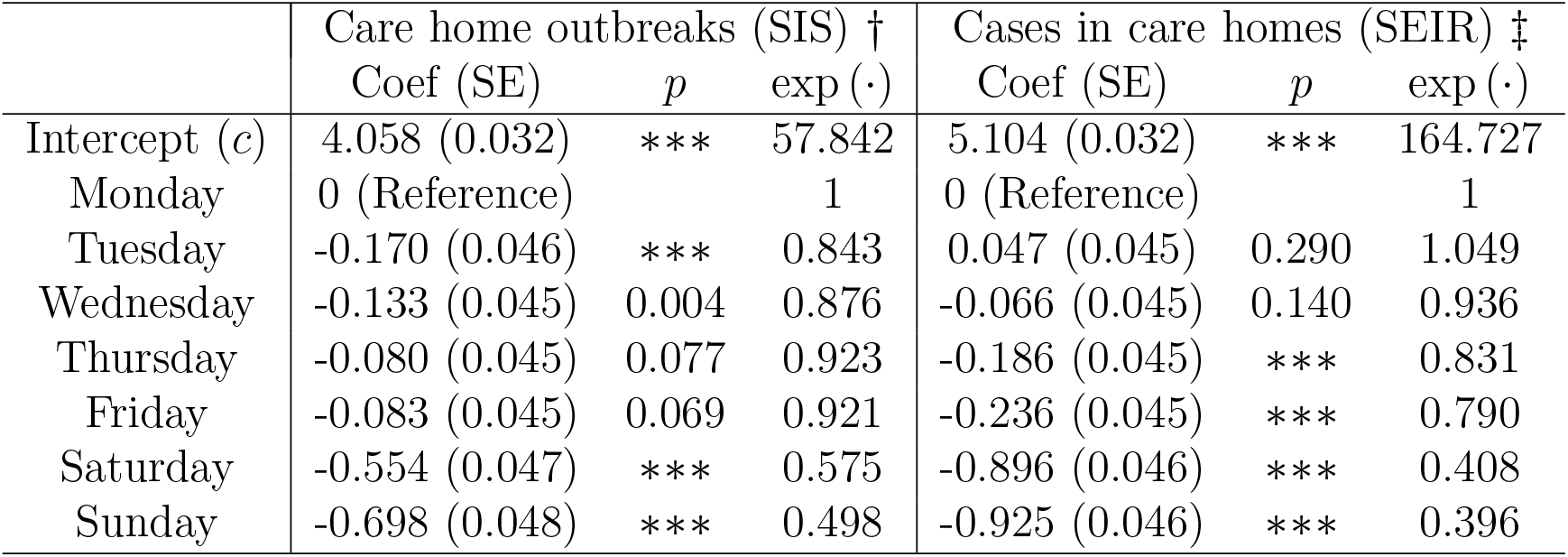
Parametric Coefficients of GAMs based on care homes experiencing out-breaks (SIS model) and individuals’ case data (SEIR model). SE is the standard error of the associated coefficient estimate (denoted Coef), the *p* column shows the resulting *p*-value and exp (·) shows the coefficient estimate on the original scale of the data. The Intercept row shows the estimate for a Monday and then the other rows show the relative difference for each day. In the *p*-value column, ‘∗∗∗’ represents coefficients with *p <* 0.001 (and so are highly significantly factors in the model). † Outbreak model: Adjusted R-squared=0.854, deviance explained=87.6%, 554 observations, smoothing parameter=14.0241 estimated by restricted maximum likelihood (REML=-2286.3). ‡Case model: Adjusted R-squared=0.92, deviance explained=95.5%, 574 observations, smoothing parameter=6.7798 estimated by restricted maximum likelihood (REML=-2937.3).

### 2.2 Modelling a time varying infection rate

The force of infection to a susceptible individual in a compartmental model is often denoted *βI/N*, where *β* is the constant infection rate per capita and *I* the total number of infectious people (at time *t*) in a population of size *N* . Rather than assuming *β* is constant we redefine this infection rate as *β*(*t*) = *νρ*(*t*), such that the time dependency is encoded in the dimensionless function *ρ* whilst *ν* is a time invariant rate defined such that *ρ* is a measure of average total infectivity spread by a single individual and that in the early epidemic phase with unconstrained exponential growth it would be equal to the usual threshold parameter *R*_0_ of an SIR-type compartmental model [4].

As such *ρ*(*t*) may be considered as a measure of the control reproduction number. Of course, the disease specific natural history such as infectious period may vary over time, particularly for extended pandemic periods over years with different variants emerging and different interventions being deployed. Thus *ν* is better thought of as simply a characteristic scaling factor and *ρ* gives insights into virological, behaviour and intervention impacts that will potentially require specific interpretation once calculated.

The infection rate *β* is a function itself of the probability of infection given contact and contact rate [7]. An intervention may appear to act to reduce the infectious period by some fraction – say isolating someone after 2 days – while in the model this effect might be rendered by keeping the infectious period the same but reducing the number of contacts. A variant may change presentation of disease and modify the infectious period (or probability of infection). The impact of variants, therefore, needs caution on interpretation of results but using a single value of *ν* allows stable interpretation of the derived *ρ* values across time.

The function *ρ* is then unknown *a priori* and our aim is to infer it from the available data, which in this work we consider being either incidence data *C*_*t*_ (new observed cases per day) or mortality data *D*_*t*_ (new deaths per day). Although the discrete data are technically the integral of the rates at which cases or deaths are observed, for simplicity we assume that the continuous functions fitted are sufficiently smoothly changing that discrete data are well approximated by the rates themselves.

#### 2.2.1 Modelling outbreak declarations in care homes

The SIS model [7] is a simple yet useful model of disease spread, and here is used to model the infectious status of entire care homes, rather than of each of the residents in them. Therefore, a care home with no reported COVID-19 positive residents is considered as a susceptible ‘individual’, which will move to the infected state when the care home detects their first case, and shall shift back to the ‘S’ stage after the outbreak is declared closed [31]. Clearly a care home cannot move to physically infect another settings but in the early stages of the pandemic staff worked in multiple care homes [32], and even without direct connection staff, visitors and residents interact with the wider community on a daily basis. Therefore the SIS structure is feasible for data showing the number of care homes currently in outbreak status, denoted by *I*, and those not in outbreak status, denoted by *S*. Then, given *S* = *N* − *I*, where *N* is the total number of English care homes (assumed constant on the timescale of a pandemic), a single equation describes the system

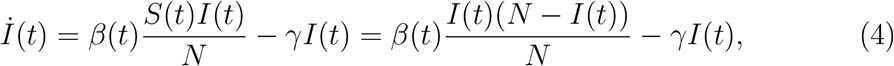

where *β*(*t*) and *γ* are, respectively, the time-varying infection rate and the removal rate. Equation (4) can also be rewritten as

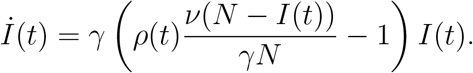

Given *R*_0_ = *β/γ* in a standard SIS model then we set *ν* = *γ* and equate the incidence measure from the GAM with the incidence predicted from the SIS model, namely

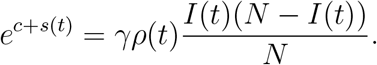

Therefore

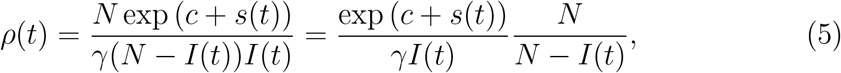

which is a measure of spread *between* care home, for example through sharing of staff or less direct transmission routes like chain of infection in members of the community. Note that, in the rightmost expression, we have factorised the control reproduction number in a term that represents the effective reproduction number and the term *N/S*(*t*), which accounts for the depletion of susceptibles in the population. Replacing the incidence *βSI/N* in (4) with *e*^*c*+*s*(*t*)^ and treating it as the external source term when applying the variation of parameters method, we obtain

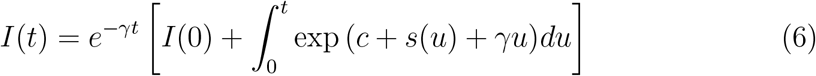

and substitution into (5) gives the following estimator of the control reproduction number directly from the time-varying growth rate *s*(*t*):

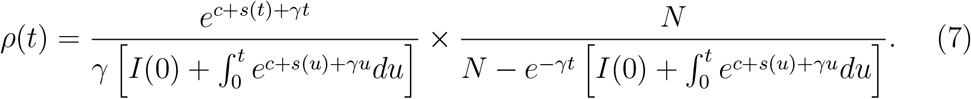

Hence, we may create an estimator of the control reproduction number *ρ*(*t*) assuming independent constant estimate values of *I*(0), *γ* and *N* . Thus, whilst there are 3 parameter values required for evaluation of the control reproduction number, we may expect *I*(0) to only be critical on short time series of data relative to the duration of a within-care-home outbreak (note that in (7) *I*(0) is scaled by a factor *e*^−*γt*^ coming from the numerator, so as *t* → ∞ the contribution of *I*(0) becomes increasingly small – as long as *s*(*t*) is not negative and too large in absolute value; see further comments on this in Appendix A.1). The parameter *N* is fixed as the observed number of care homes in England and is derived from the Care Quality Commission. Hence the parameter *γ* is likely the most critical and sensitive one. We can take *γ* = 1*/T* where *T* is the average duration of an outbreak. In this case, for a point estimate of *ρ*(*t*) at a fixed time *t* we may sample estimates of *c* and *s*(*t*) from surveillance data and include uncertainty on *T* by sampling from the distribution on mean outbreak duration assuming *N* is fixed and *I*(0) has burnt off. For simplicity, however, in this work we fix *T* so credible intervals reflect only the uncertainty from the GAM fit to surveillance data.

The effective reproduction number, denoted *R*_*E*_(*t*), in this case is

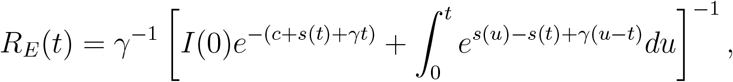

which is obtained from (7) by simply writing the first term in an alternative form and disregarding the second term, which captures the depletion of susceptibles. Note that, in a large population with low initial prevalence, the depletion of susceptible is negligible for some time, so control and effective reproduction numbers are approximately equal, i.e. *ρ*(*t*) ≈ *R*_*E*_(*t*). In Appendix A.1 we show that, in this case, an even simpler approximation to both reproduction numbers 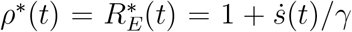 can be derived when assuming any transients arising from the initial conditions have passed and *s*(*t*) is approximately linear, i.e. the incidence is growing or declining approximately exponentially, for some time.

#### 2.2.2 Modelling individual cases across care home settings

For data measuring case incidence, an SEIR model structure is assumed as a representative approximation to the disease dynamics. The standard SEIR model might need some adaptation for application to a residential setting (such as care home or prison) as some of the residents might pass away or leave due to non-COVID reasons, and then the arising vacated rooms would be filled by new residents. For simplicity, we modelled this replacement as instantaneous (though in reality there may be a period of vacancy), but we allow new individuals to be either susceptible or immune (e.g. both due to natural infection or vaccination). Therefore, we have

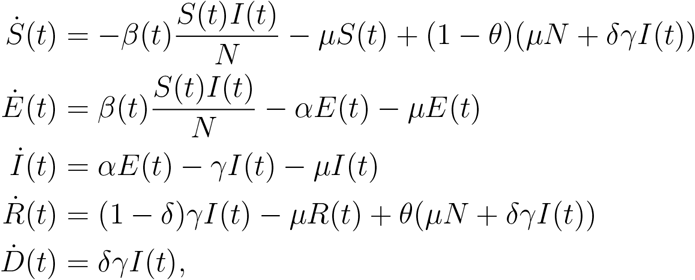

where disease states *S, E, I, R* and *D* denote the numbers of susceptible, exposed, infected, recovered and deceased, respectively, *β*(*t*) and *γ* are (as before), the infection and removal rate, respectively, *α* is the rate of becoming infectious, *δ* is the COVID-19-specific case fatality ratio, *μ* is the rate at which residents leave the care home (natural mortality rate) and *θ* is the proportion of new entries that have immunity to the infection (this is not necessarily the same as conferred immunity in residents and staff from past outbreaks). An individual leaving the population is assumed to be immediately replaced by new individual (immune with probability *θ* or otherwise susceptible), so the total population *N* = *S* +*E* +*I* +*R* (i.e. the sum of those in living state) is constant, and *D* is effectively only used to keep track of the number of COVID-19 deaths. In the use case that the surveillance data does not permit tracking the population turnover due to natural mortality, *μ* may be set to zero. Note that *θ* will in general vary with time due to vaccination policy or community epidemics, but at the start of a new pandemic we expect *θ* = 0.

To derive an estimator for the time varying reproduction number *ρ*(*t*) we substitute *β*(*t*) = *νρ*(*t*). In this model disease incidence (new entrants to *I* state) is given by *αE*(*t*) = *e*^*c*+*s*(*t*)^. Solving for *E, I* and *R* means that

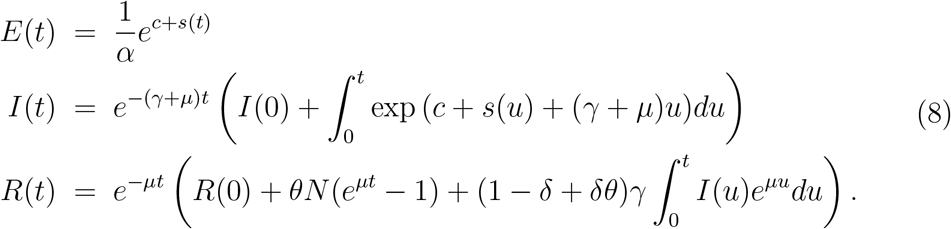

We are thus assuming that *E*(0) = *e*^*c*+*s*(0)^*/α*; other initial conditions (*I*(0) and *R*(0)) are discussed below.

Because individuals can leave the care homes due to natural mortality, the average time length of any infected individual being infectious is 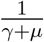 rather than 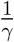, and individuals leave the *E* stage at the rate of *α* + *μ* but the rate of entering the *I* stage is *α*. Therefore, only a fraction 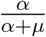 of the infected individuals can become infectious, with the opposite fraction dying while in the *E* state. Thus, in this model the critical scaling timescale on the reproduction number is

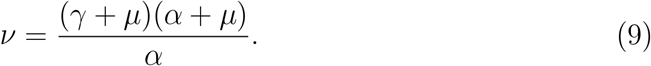

The control reproduction number estimator is given by

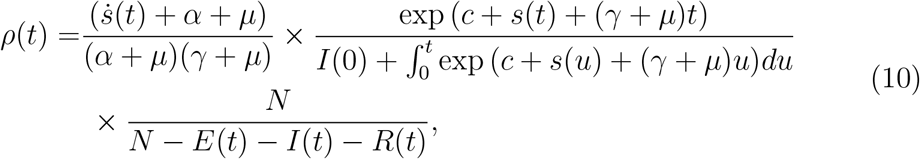

where *E*(*t*), *I*(*t*) and *R*(*t*) are as in (8). The estimator *ρ*(*t*) may then be inferred from surveillance data for independent estimates of parameters *γ, α, μ, δ, θ, I*(0), *R*(0) and *N* . As in the previous section, the initial conditions (*I*(0) and *R*(0)) have only transient effects and population size *N* can be learnt from the surveillance data being used. In this study we assume for simplicity low community immunity and limited community vaccination, and hence *θ* = 0. However, future work will consider the impact of vaccination (community and setting specific) and a time-varying *θ* on calculation of the control reproduction number. In the main application in Section 3.3 we will also further assume for simplicity no natural or disease-induced mortality (*μ* = *δ* = 0 – for a case where *δ >* 0, see Appendix A.2). The disease natural history parameters are then likely the most sensitive parameters to estimate independently.

The effective reproduction number is then obtained when the last factor *N/S* in (10) is removed:

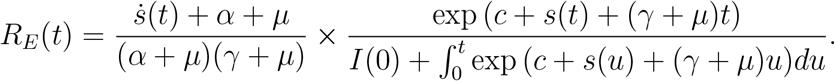

When the population is almost entirely susceptible (prevalence, incidence and *θ* are small), the effective and control reproduction numbers are approximately the same. In this case, with similar arguments as in Appendix A.1, assuming the transient has burnt out, that replenishment is slower relative to disease transit rates (*μ* is negligible compared to *γ*), and that the incidence is growing approximately exponentially for a while, we may derive a similar approximation to that for the SIS model (compare with [15]),

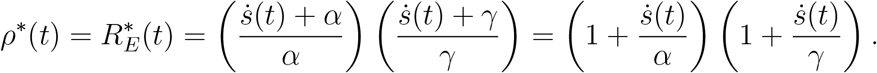

The methods presented here can be adapted to consider mortality data rather than case data: an application of this model version to simulated data is provided in Appendix A.2.

### 2.3 Calibration on simulated data

To check if the estimators derived above are appropriate and useful, a first test is to apply them to stochastically simulated data from the epidemic models described above. Keeling and Rohani [33] illustrate the application of the Gillespie algorithm to such epidemic models [34]. Event-based algorithms are typically slow and so a ‘*τ* -leap’ method [35] is used here instead.

The SIS simulation (Figure 1, top panel, showing the arising incidence from the SIS model) is initiated with population *N* = 10^4^ and *I*(0) = 100, *γ* = 0.05 (20 days for recovery on average), both chosen as indicative of the number of care homes and duration of outbreaks, with a time step of Δ*t* = 0.1 days. The transmission rate is taken to be *β* = 0.5 for *t <* 12 and is assumed to drop to *β* = 0.25 for *t* ≥ 12 to simulate the impact of an intervention at time *t* = 12. This gives *R*_*C*_ = *R*_0_ = 10 before the intervention, with a control reproduction number that drops post intervention to *R*_C_ = 5. We note that *R*_0_ = 10 is large and so spread is rapid, but the value is not excessive given the interpretation is that this represents the average number of care homes infected by a single care home over its entire outbreak. An SIS model should have an endemic equilibrium predicted by 1 − 1*/R*_0_ and hence we expect the incidence in this simulation to approach 400 care homes in outbreak state as seen by end of the simulation. Figure 1 top panel also shows the central estimate from the GAM fitted to this data.

**Figure 1:**
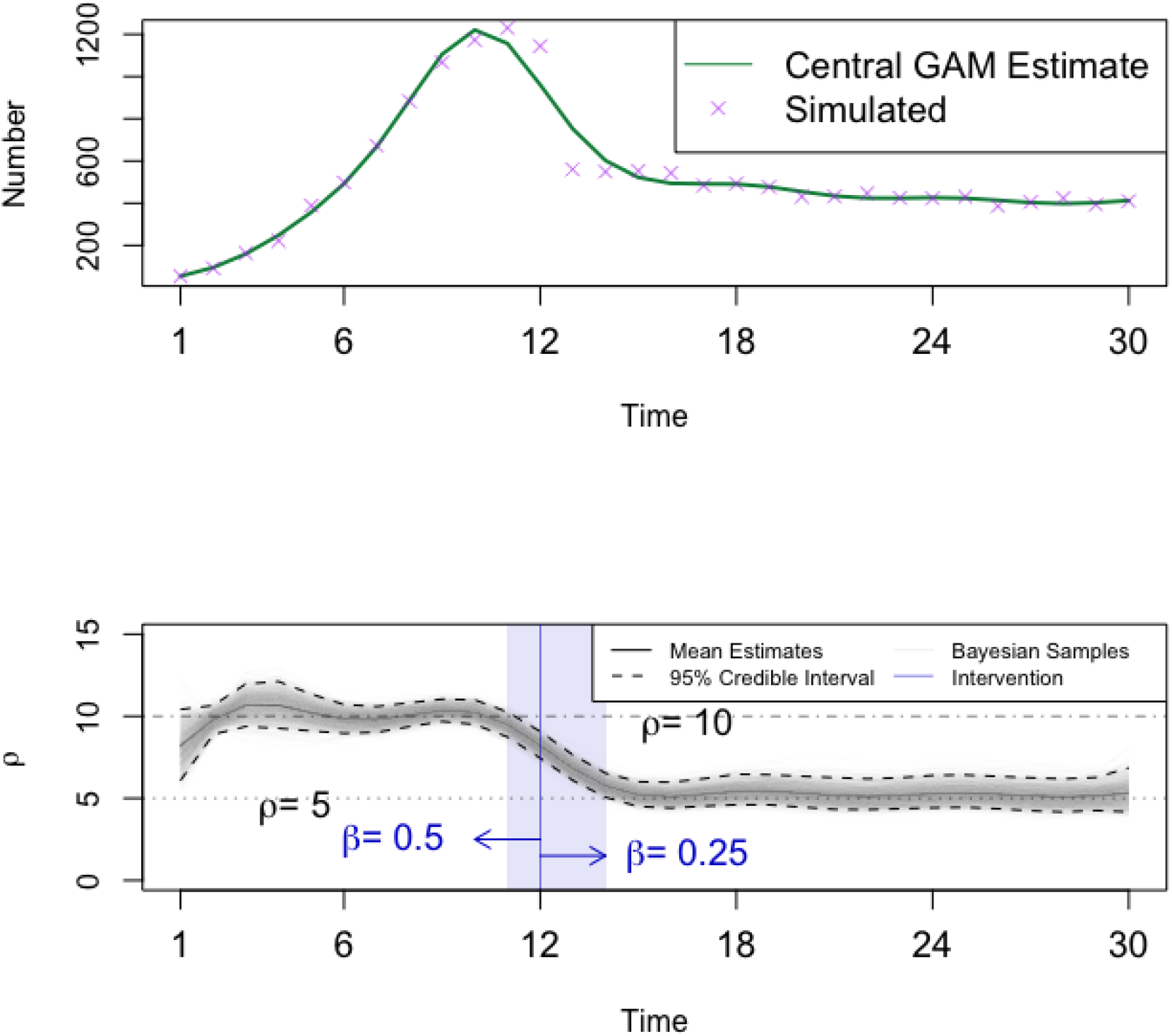
Top: Stochastic simulation (*τ* -leap) for the SIS model. Parameter values are: *N* = 10^4^, *I*(0) = 100, *β* = 0.5 → 0.25 at day 12, *γ* = 0.05, Δ*t* = 0.1. Purple crosses represent the simulated incidence data (number of new individuals entering state *I* per day) and the green line the resulting GAM fit, assuming `family=quasipoisson`. Bottom: Estimated reproduction number *ρ*(*t*) (black solid line) from the simulated data from the SIS model. The Bayesian sample (light gray lines) is generated using the conditional simulation method in [28] and from the sample the 95% CrI (black dashed lines) is obtained. The intervention *β* = 0.5 → 0.25 at day 12 is depicted by the blue vertical line. There is no transient time, while the adjustment time is presented by the blue shaded band.

The SEIR model (Figure 2, top panel, showing the arising incidence from the SEIR model) is simulated similarly and initialised with 50 infected and 50 exposed individuals among a total population of 10^4^. The average incubation period is 7 days (*α* = 0.143), the average infectious period is 10 days (*γ* = 0.1), and the time step used in the simulation is Δ*t* = 0.15 days. For simplicity, we set the rate at which residents leave to care home *μ* = 0, the community immunity *θ* = 0 and the case fatality ratio *δ* = 0. To model an intervention at time *t* = 90, the infection rate is defined as *β* = 0.143 for *t <* 90, and *β* = 0.1 afterwards. The uncontrolled outbreak will thus have *R*_0_ = 1.43 whilst, post-intervention, *R*_C_ = 1 (see dashed-dotted and dotted lines in bottom panel of Figure 2, respectively). There is no assumed reporting bias by weekday. The incidence peaks close to time of the intervention that reduces *R*_*C*_ to 1, but with the built-up residual immunity the effective reproduction number is below 1 and hence the epidemic declines. Figure 2, top panel, also shows the derived GAM central estimate.

**Figure 2:**
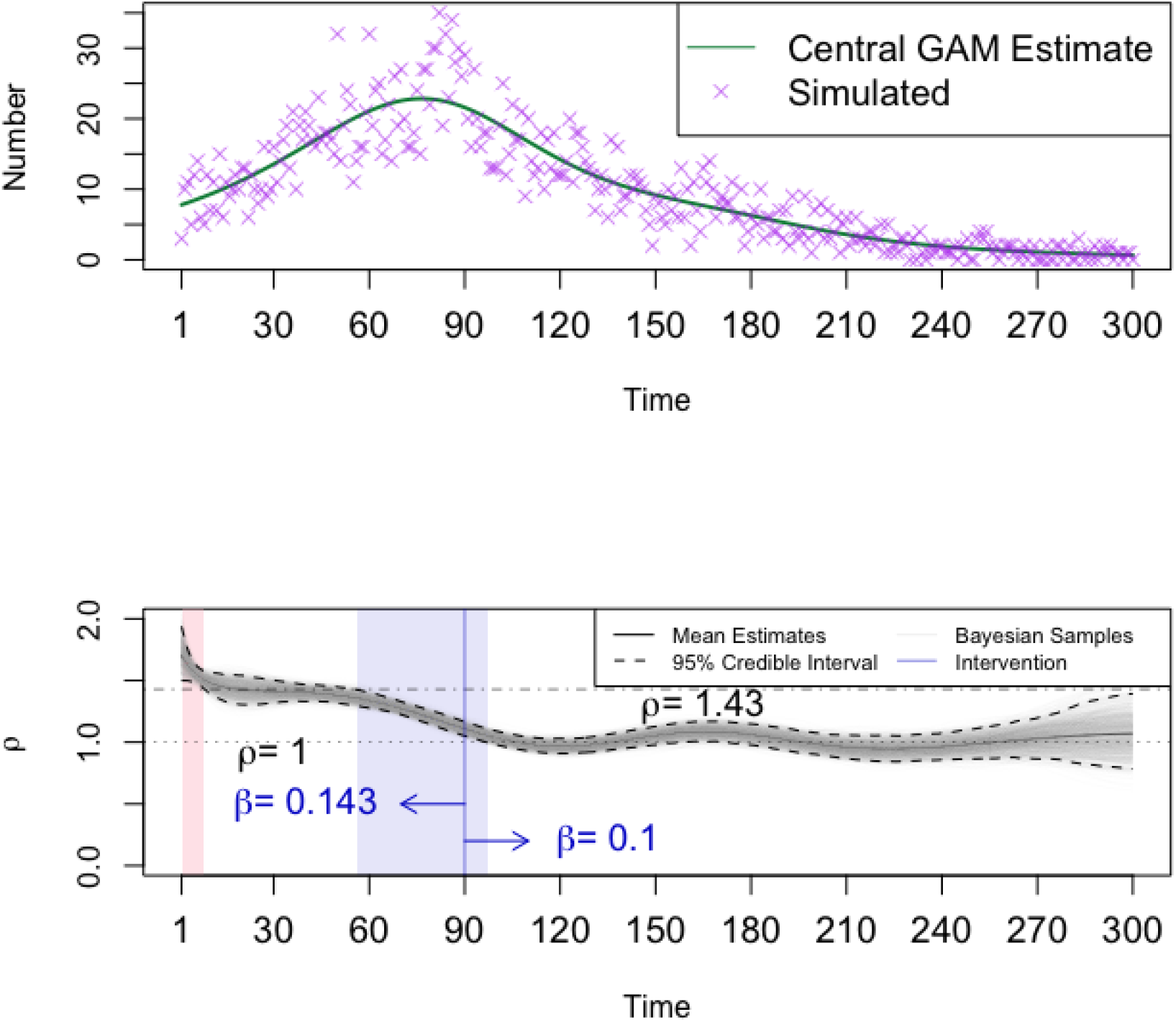
Top: Stochastic simulation (*τ* -leap) for the SEIR model. Parameter values are: *N* = 10^4^, *E*(0) = *I*(0) = 50, *β* = 1*/*7 → 0.1 at day 90, *α* = 1*/*7, *γ* = 0.1, Δ*t* = 0.15. Purple crosses represent the simulated incidence data (number of new individuals entering state *I* per day) and the green line the resulting GAM fit, assuming `family=quasipoisson`. Note that, compared to Figure 1, the much more pronounced natural stochastic variability in the data is purely due to the much longer epidemic and the much lower daily incidence (i.e. no observational error has been added to the simulation output). Bottom: Estimated reproduction number *ρ*(*t*) (black solid line) from the simulated data from the SEIR model. The Bayesian sample (light grey lines) is generated using the conditional simulation method in [28] and from the sample the 95% CrI (black dashed lines) is obtained. The intervention *β* = 1*/*7 → 0.1 at day 90 is depicted with the blue vertical line. The transient and adjustment time are depicted by pink and blue shaded bands, respectively.

### 2.4 Data

Having tested that the method is able to recover known parameters from simulated data, the method is the applied to real world data sets.

All positive COVID-19 tests were asked for the residential address, though when not available the GP-registered address was used. Within UKHSA, address information was provided by the Second Generation Surveillance System (SGSS) [36] and the UKHSA Geospatial Team then matched addresses to the Ordnance Survey database to obtain a Unique Property Reference Number (UPRN) and Basic Land Property Unit (BLPU) class information.

Addresses with BLPU classes of RI01 were identified as care homes. Care Quality Commission (CQC) IDs for care homes and their accompanying UPRN information were also linked to the cases line list to identify registered care providers not identified through the address matching process. Finally, properties with ‘care home’, ‘rest home’, ‘senior living’, ‘elderly’, ‘retirement’ were also searched to determine whether there were any additional addresses that might have been care homes [37, 38]. The total number of care homes for older people is about 14,500, with a total of about 450,000 beds associated with these settings [39]. We use the number of beds as a proxy for the population size. We expect this to be an overestimate as care homes typically operate at 90% capacity, but a precise value of the number of residents is hard to specify and likely fluctuates significantly over time.

This linked dataset was used to create two aggregated and non-identifiable samples, namely:

1. Outbreak declaration data: the number of care homes in England that declared an outbreak on given days from 01/04/2020 to 06/10/2021.
2. Test-Positive data: all cases attributed to adult social care settings were aggregated by day of positive test to give the number of new confirmed cases among care home residents in England from 15/03/2020 to 09/10/2021.

The estimated time-varying reproduction numbers inferred are relevant to the data set (and underlying population) in question and may differ from published national estimates for other settings or communities. Furthermore we do not consider different risk group reproduction numbers, as this would require, for example, separate data on residents and staff in homes. Such structured analysis is not possible with current data and is subject to future research. As such, the estimated reproduction number can be considered indicative of the (additional) control effort required at that time but may not formally translate to point estimates of transmission from specific settings.

## 3 Results

### 3.1 Simulation results

In the numerical result shown in Figure 1, bottom panel, the estimation of reproduction number is roughly 10 before day 11, and 5 after day 14. Remember that in the simulation we set the infection rate to drop on day 12 (blue line), so the estimator *ρ* appears to decrease before the intervention, likely because of interplay between the stochastic noise and the number of knots used in model, as the GAM tries to smooth through the data points.

For clarity, it is worth defining the concepts of transient and adjustment times in simulation results as follows:

- Transient time: the period, at the beginning of the simulation, during which the credible interval does not include the pre-intervention reproduction number.
- Adjustment time: the period from the time the upper credible interval stops including the pre-intervention *R*_0_ to the time when the lower bound firstly includes the post intervention *R*_0_.

It may be possible for either or both time periods to have length 0 in some simulations.

There is no transient time evident for the SIS simulations (see Figure 1, bottom panel, and 3), but the adjustment time is present regardless of the choice of parameters. In Figure 3, (top left panel, when *N* = 10^3^, *I*(0) = 100), the adjustment time is very short (the window is effectively compressed to a single day, which occurs after the simulated change event), likely because of the relatively small population and large initial case numbers; when *N* = 10^4^ and *I*(0) = 10 (top right), the adjustment is slower and includes the actual time when the parameter changed in the simulation; a similar situation occurs when *N* = 10^5^ and *I*(0) = 100 (bottom), with an adjustment time that appears very similar in the cases of an early (bottom left) or late (bottom right) change time. The top left results suggests that caution may be needed in applications with very high initial case numbers and high transmission.

**Figure 3:**
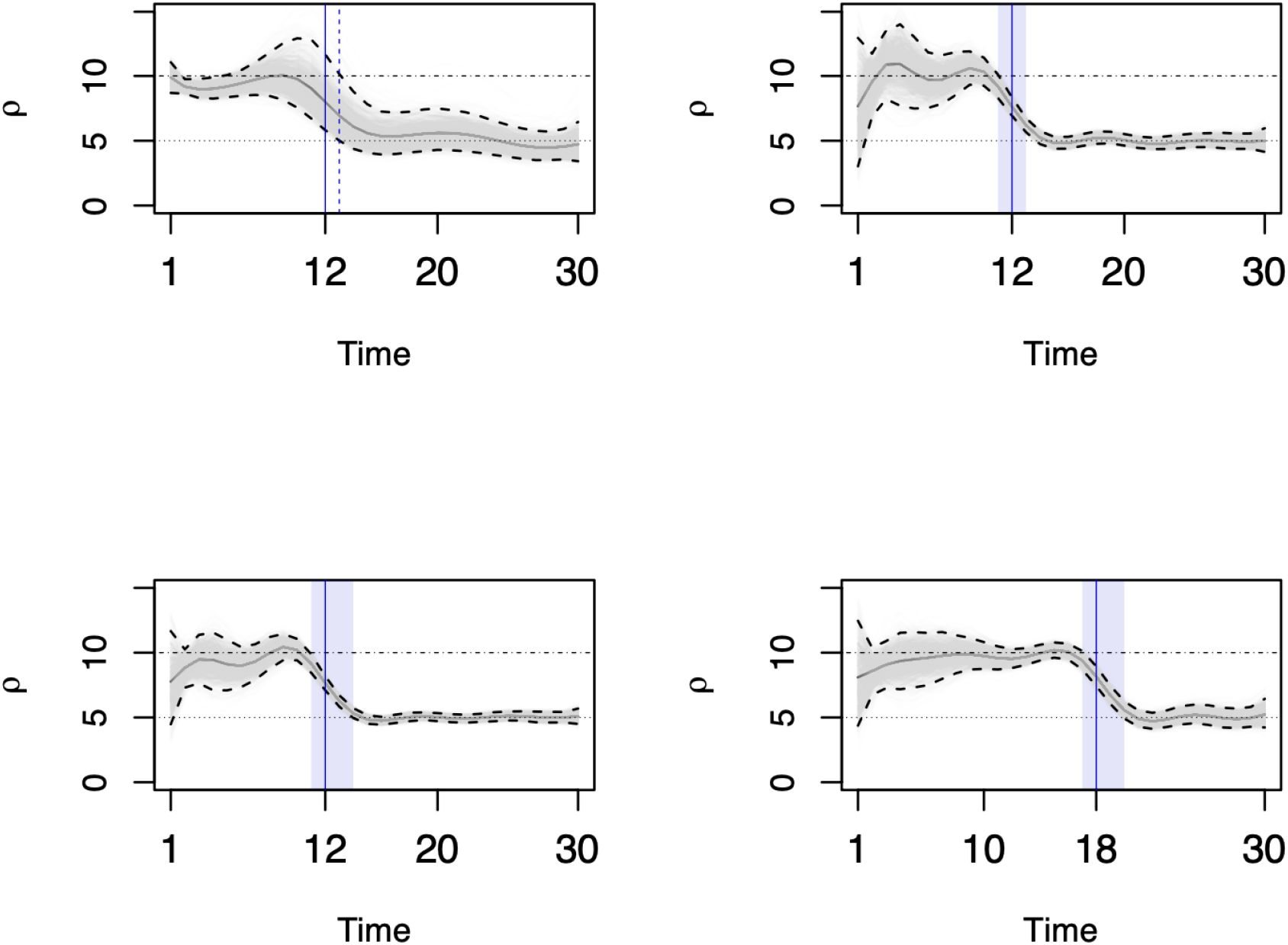
Estimated reproduction number for the stochastic SIS model for other parameter choices. Top left: *N* = 10^3^, *I*(0) = 100; top right: *N* = 10^4^, *I*(0) = 10; bottom left: *N* = 10^5^, *I*(0) = 100; bottom right: *N* = 10^5^, *I*(0) = 100 and intervention at day 18. In all plots, *γ* = 0.05. The Bayesian sample (light gray lines) is generated using the conditional simulation method in [28] and from the sample the 95% CrI (black dashed lines) is obtained. The intervention *β* = 0.5 → 0.25 is depicted by the blue vertical line. The transient period does not occur, while the adjustment time is depicted by a vertical dashed line (top left) and blue shaded bands (other panels).

The estimation results for the SEIR simulations (Figure 2, bottom panel) follow a similar pattern to the results obtained from the SIS cases, except that there appears to be a transient phase (likely due to the characteristic polynomial being quadratic – unlike in the SIS model case – and therefore the proportions of cases in *E* and *I* taking some time to converge to the dominant eigenvector of the matrix describing the linearised system). As for the SIS model, the estimated reproduction number decreases around the sudden change in transmission, although this time more gradually (Figure 2, bottom panel). The *E* and *I* compartment numbers drop to relatively low levels (similar to that of the epidemic initial conditions), from about day 150 onwards (not shown), and keep going down afterwards, resulting in decreasing size of daily new cases and hence increasing estimation uncertainty as shown by the widening credible interval at late times.

In Figure 4, the transient time is usually 7 to 12 days while the adjustment time is mostly around 30 days. The transient time is short relatively to the whole time span of epidemic wave. The adjustment time started 20 to 30 days ahead of the intervention day, and includes the actual intervention time. When interpreting the results of this method in a real world application, this adjustment phase may require consideration (though in the real world an intervention is unlikely to have a step function impact, instead rather exhibiting variation in uptake and effectiveness both in time and space).

**Figure 4:**
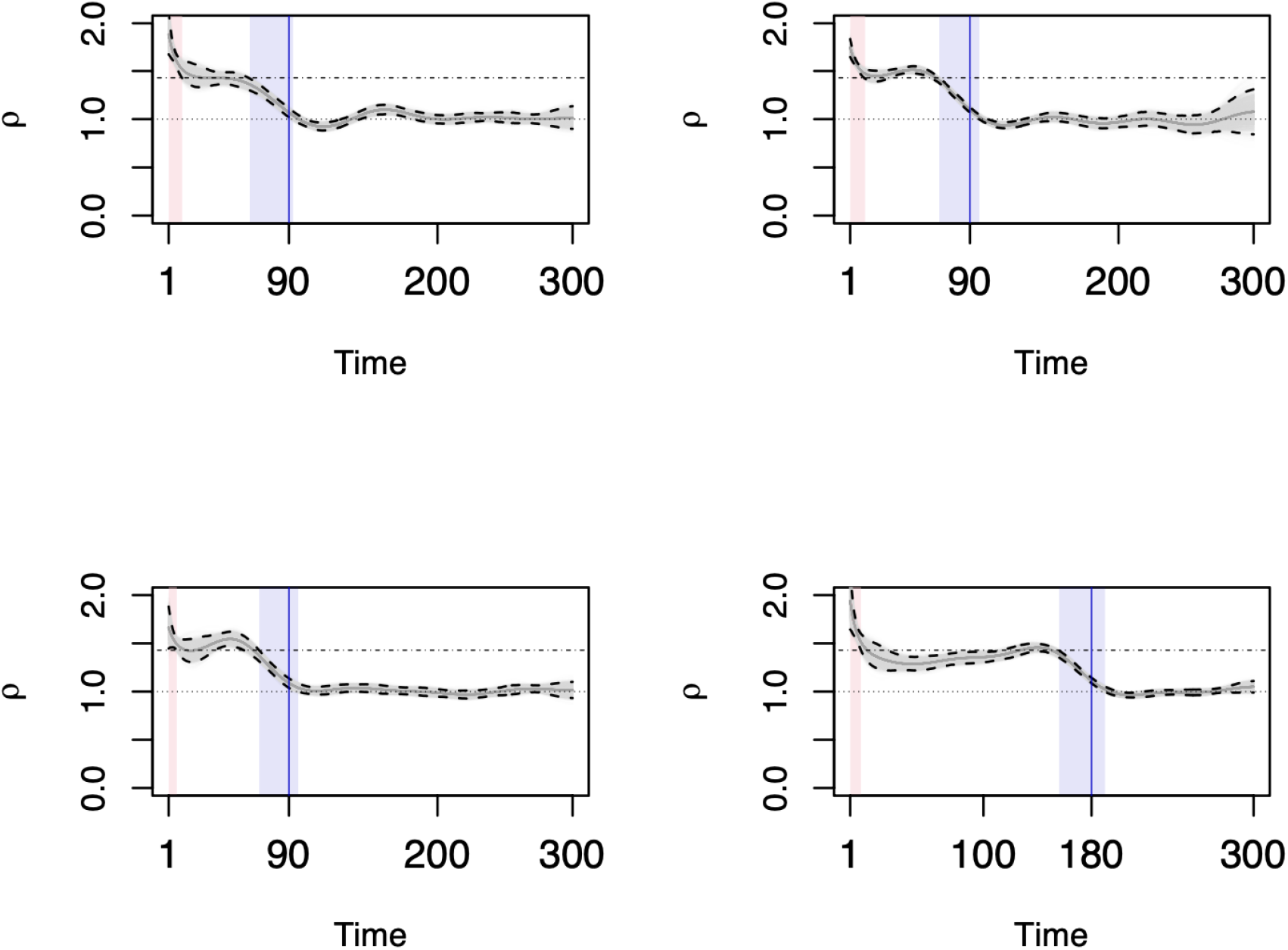
Estimated reproduction number for the stochastic SEIR model for other parameter choices. Top left: *N* = 10^5^, *E*(0) = 50, *I*(0) = 50; top right: *N* = 10^5^, *E*(0) = 500, *I*(0) = 500; bottom left: *N* = 10^6^, *E*(0) = 50, *I*(0) = 50; bottom right: *N* = 10^6^, *E*(0) = 50, *I*(0) = 50, intervention at day 180. In all plots, *γ* = 0.05. The Bayesian sample (light grey lines) is generated using the conditional simulation method in [28] and from the sample the 95% CrI (black dashed lines) is obtained, and the intervention *β* = 1*/*7 → 0.1 is depicted by the blue vertical line. The transient and adjustment time are depicted by pink and blue shaded bands, respectively.

### 3.2 SIS: outbreak data

Stable reporting of outbreaks started in April 2020. According to the outbreak data, about 150 to 200 care homes per day started experiencing an outbreak during April of 2020, although this number then dropped in the following three months. After the summer holidays in July and August 2020, the outbreak counts climbed up again until the second national lockdown in November 2020. However, a higher peak at 250 was reached during the Christmas break, followed by a dramatic decrease during the period of the third lock down and the start of the vaccine campaign from January to March of 2021. The spread of SARS-CoV-2 in the English care homes then apparently eased for two month until June of 2021. After that, a new wave of infection then took place again when the ‘Delta’ variant started to spread around the country, although the daily counts stayed mostly under 100 (Figure 5, top panel, black line).

**Figure 5:**
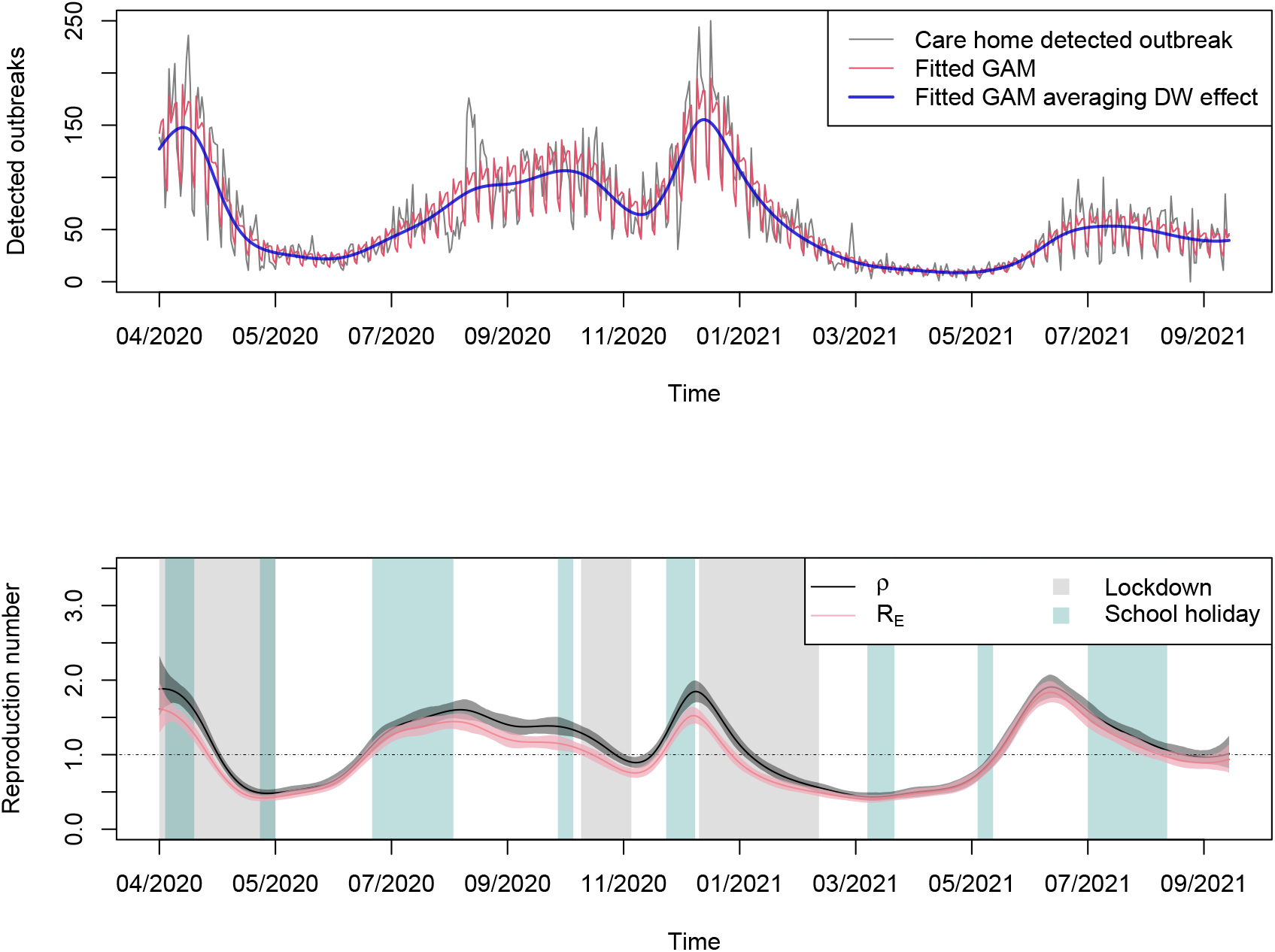
Top: GAM results with options `family=nb(link=log), bs=‘ps’` and `k=27` knots for data on English care home experiencing an outbreak from 01/04/2020 to 06/10/2021. The data (black line) shows small spikes every 7 days, which are captured as day-of-the-week (DW) effects and interpreted by parametric coefficients (see Table 1 for estimates) of the GAM fit (red line). The DW effects can be removed (blue line) by taking an average of the 7 values (using 0 for Monday as reference day). Bottom: Mean estimate (line) and 95% conditional credible interval (shaded band) for *ρ*(*t*) (black) and *R*_*E*_(*t*) (pink) for the English care home outbreak data (SIS, *N* = 14, 500, *I*(0) = 2, 000, *γ* = 1*/*26). The control reproduction number *ρ*(*t*) and the effective reproduction number *R*_*E*_(*t*) are generally observed to decreases during lockdowns (grey shaded bands) and increase during the holidays (green shaded bands), at least before widespread vaccination in care homes at the start of 2021. A dotted horizontal line is added to highlight when the estimated reproduction numbers cross the unity threshold.

The outbreak data shows periodic weekly fluctuations, indicating the presence of day-of-week effects in the reporting mechanism. Therefore, the generalised additive model (GAM) fitted to outbreak data should include a parametric term for day-of-week effects (see (3)). The smoother uses the penalised spline and the number of knots is set to 27 for 554 days in total (`bs=‘ps’, k=27`).

The coefficients of intercept and factors (weekdays) are summarised in Table 1, referring to ‘Monday’ as the baseline. Overall, Monday usually sees the most reported outbreaks, while Tuesday and Wednesday’s amount are decreased by 1*/*6 and 1*/*8 on average, respectively, compared to Monday. Thursday and Friday (with *p*-value being not significant at a 5% level), both expect numbers of reported outbreaks closer to those of Monday. At the weekend, the average count of reported outbreaks are roughly half of Monday’s. Therefore taking arithmetic mean of the coefficients of ‘weekday’, with ‘Monday’ as 0, can help remove the ‘weekly noise’ and produce a smooth representative average curve (Figure 5, top panel, blue line) for integration with the compartmental model.

In Figure 5, bottom panel, green shaded bands identify the Christmas break, Easter break, summer holiday and half terms, times during which most adults and students are on holiday, and care homes may have more visitors than usual. The grey shaded bands stands for the three main lockdowns that aimed at reducing the disease transmission nationally and therefore reduced the interaction between different care homes and between care homes and the community. Before March 2021, *ρ* decreased during the lockdown periods and then increased during the summer period when restrictions relaxed, indicating that the governmental intervention of reducing social community contacts has likely been helpful for care homes’ epidemic prevention. We note that testing became more widespread from late Summer 2020, so the increase from August may be an artefact of historical cases being detected from Polymerase Chain Reaction (PCR, which tests for RNA rather than live virus) and hence may result in an apparent boosting the number of outbreaks.

### 3.3 SEIR: case incidence data

The case incidence data (technically, the number of new individuals testing positive to infection) is taken as a proxy for the number of daily new residents becoming infectious in care homes throughout England. The timeline for these case data is 15/3/2020 to 09/10/2021 (Figure 6, top panel), with a similar curve trend as that observed for the outbreak data above (Figure 5, top panel). Although this time series covers about a year and a half of data, which is roughly the same as the average lifespan in care homes due to natural mortality, for simplicity and to highlight the difference between control and effective reproduction number, we assume negligible population turnover due to both natural and disease induced mortality and negligible immunity in the population in the model in Section 2.2.2 (i.e. *μ* = *δ* = *θ* = 0 – see instead Appendix A.2 for a model with *δ >* 0), thus effectively turning it into a standard SEIR model. The first epidemic peak of care home cases happens at the end of April, with over 1000 diagnoses on some days, and a double peak between October 2020 and February 2021 reaching values of about 1400 and 2500, respectively. After the release of the third lockdown, a much smaller sized epidemic is seen when the new variant ‘Delta’ appeared in mid-2021 (Figure 6, top panel, black line).

**Figure 6:**
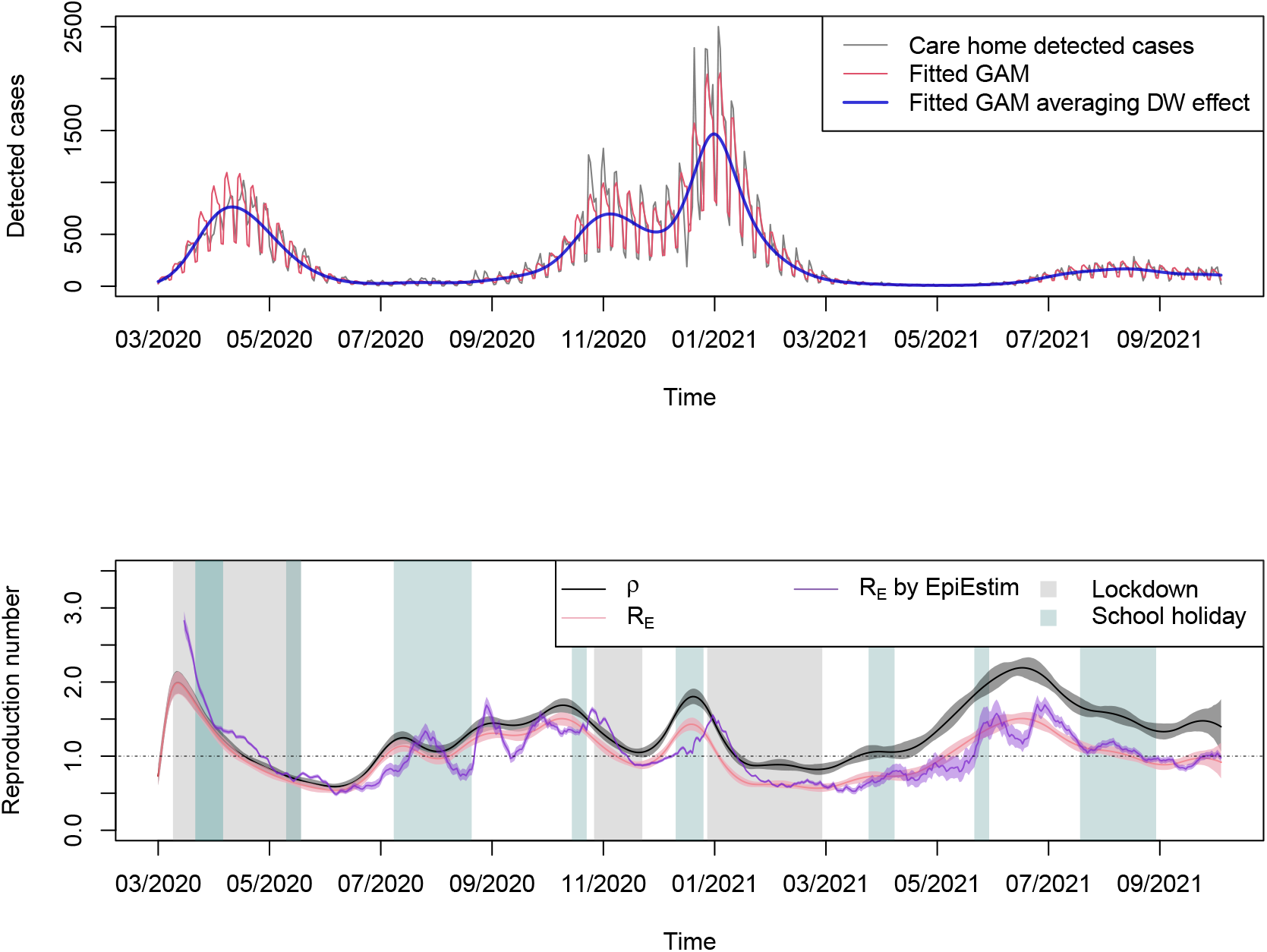
Top: GAM results with options `family=nb(link=log),bs=‘ps’` and `k=28` for the case data recorded in English care homes from 15/03/2020 to 09/10/2021. The data (black line) shows small spikes every 7 days, which are captured as day-of-the-week (DW) effects and interpreted by the parametric coefficients (see Table 1 for estimates) of the GAM fit (red line). The DW effects can be removed (blue line) by taking an average of the 7 values (using 0 for Monday as reference day). Bottom: Mean estimate (line) and 95% conditional credible interval (shaded band) for *ρ*(*t*) (black) and *R*_*E*_(*t*) (pink) for the case data from English care homes (SEIR, *N* = 450000, *I*(0) = 500, *R*(0) = 0, *γ* = 1*/*5, *α* = 1*/*3). The control reproduction number *ρ*(*t*) and the effective reproduction number *R*_*E*_(*t*) are generally observed to decrease during lockdowns (grey shaded bands) and increase during the holidays (green shaded bands), at least before widespread vaccination in care homes at the start of 2021. The mean estimate (purple line) and 95% credible interval (purple shaded band) from `EpiEstim` (with serial interval following a Gamma distribution with mean 8 and standard deviation 5.83 days) follows the general scale and trend of the effective reproduction number estimated with the method proposed here. A dotted horizontal line is added to highlight when the estimated reproduction numbers cross the unity threshold.

As for the outbreak data, the case data also displays day-of-the-week effects. The case data is fitted with a GAM (3) with penalised spline as smooth term and 28 knots for 574 days in total (`bs=‘ps’, k=28`), and the fitting results are presented as red and blue lines in Figure 6 (top panel). The parametric coefficients are summarised in Table 1, with Monday as baseline for the day-of-the-week effect. During the week, the numbers of cases for Tuesday and Wednesday are not significantly different from those on Monday, while the newly positive case counts for Thursday and Friday are about 80% of those observed on Monday. Weekends are expected to have fewer positive testing results, with both Saturday and Sunday having about 40% of Monday’s cases. This likely reflects the average operational timing of regular testing in such settings.

Figure 6 (bottom panel) shows the reproduction number and its credible interval after averaging the day-of-the-week effects. The results have a similar pattern as seen in Figure 5 (bottom panel), with the three national lockdowns corresponding to periods of reducing values of the reproduction numbers. The control reproduction number (black line and dark grey shaded band) reaches values higher than 2.5 in March 2020, increases from 1 to 1.5 during summer of 2020, and peaks at a value close to 2 in October and December 2020. From December 2020 care homes received the vaccine and so the fast change from *ρ* = 2 to *ρ* = 0.5 is likely a combination of community lockdown and vaccine protection within settings. In 2021, the reproduction number peaks between June and July, when the ‘Delta’ variant was the dominant variant in England [40].

### 3.4 Comparing with EpiEstim

The `EpiEstim` R package has been developed to compute the effective reproduction number by combining the incidence data with information from the serial interval, which can either be assumed to follow a plausible prior or be directly obtained from some surveillance system [11, 13]. We compare `EpiEstim` to the results obtained with the method presented here when both are applied to the case data from English care homes (Figure 6, bottom panel, purple and pink lines and shaded bands). In the parametrisation of the SEIR model the average incubation period is set to 3 days and the average infectious period to 5 days. These are plausible values for SARS-CoV-2 (e.g. in line with [41]) and are taken at face value (i.e. in particular, without any uncertainty) given we are only seeking an indicative parametrisation to illustrate the methodology rather than focusing on a precise characterisation of the timing of disease progression. For this choice of parameters, the generation time distribution is right-skewed, with mean 8 days and standard deviation of 5.83, and with probability density function [15, 42]

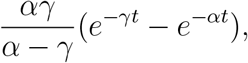

which is close to a Gamma distribution with the same mean and standard deviation. We input this Gamma as the serial interval in `EpiEstim`, implicitly assuming that the generation time is equivalent to the serial interval, and use a sliding window of 14 days. The comparison between `EpiEstim` and the estimate obtained with the methodology proposed here for the SEIR model is presented in the bottom panel of Figure 6 (purple and pink lines and shaded bands). Although the results of the two methods show some differences, which is to be expected given that the approaches are substantially different (e.g., `EpiEstim` does not account for a day-of-the-week effect, which likely leads to the shorter time-scale oscillations compared to our method), the results show a similar general scale and trend.

## 4 Discussion

This work brings together semi-parameteric generalised additive modelling (GAM) with traditional structured compartmental epidemic models. The GAM can be used to calculate an instantaneous growth rate of time series data and has an advantage that it can parameterically incorporate day-of-the-week effects to avoid unnecessary aggregation (e.g. to weekly numbers or rolling averages) to work around reporting effects. Layering on top of the GAM mechanistic assumptions about disease epidemiology, we have shown we can generate an estimate for the control reproduction number – the number of people infected per case in wholly susceptible population, but with mixing patterns modified by a control policy or spontaneous response to an outbreak – and the effective reproduction number – the number of people infected per case given current mixing patters and, importantly, partial susceptibility in the population.

This method for calculating time-varying control and effective reproduction numbers compares well with the alternative approach given by `EpiEstim` and is verified by simulation studies. Application to real world data is promising. However, care homes are home to elderly people with strong staff contacts and it is not clear what impact age has on the mechanics of disease progression. Therefore, these estimates are mainly illustrative of the methodology and further work is needed to compare them with the transmission estimates from community surveillance. The method shown here is applied to data about active outbreaks and case data, but it can be adapted to mortality data (see Appendix A.2).

Limitations include not incorporating time-varying case fatality ratio *δ* and community immunity *θ* that in reality is known to have changed during the pandemic. However, all methods are sensitive to these changes and by parametrically including these effects one can be explicit in the assumptions for future interrogation. A second limitation is that single type compartmental models assume homogeneous mixing in the population which means the method may be biased if the outbreak had core groups sustaining transmission. This is also true of other established tools such as `EpiEstim`.

At present, the uncertainty reflects only the inherent uncertainty from the GAM rather than uncertainty associated to the compartmental model parameter estimates. Furthermore, the SEIR model used assumes Markovian transitions between stages. This is a simplification but, as stated above, the intention is to get a representative metric for instantaneous transmission to aid decision makers’ thinking. Other model structures may be included at the cost of increased complexity in computation and understanding.

The method requires the specification of an initial condition *I*(0). The choice of *I*(0) affects the estimate of *ρ* towards the beginning of the time series, though its impact wanes on the timescale of a generation time. However, during this transient phase, the estimate of *ρ* may need careful interpretation. The robustness of the results could be improved by incorporating uncertainty in *I*(0) as part of the inference scheme developed here or allowing the estimate of *ρ* to be presented only after a time period comparable to the generation time has elapsed (as is the case with `EpiEstim`).

Similarly to *I*(0), results also depend on the choice of the initial condition for the immune population *R*(0). The impact of this choice also wanes over time, but affects *ρ* during these transients. However, given the pandemic context, it is reasonable to set *R*(0) = 0 and let the immunity build up as more cases get infected. For simplicity, we are accepting the data as an accurate reflection of the number of individuals infected in care homes over entire time series. However, we acknowledge that, due to low case ascertainment in the first wave, there may have been a larger depletion of susceptible population than we have accounted for here. Estimates of the effective reproduction number are not affected, but an underestimation of the immune population translates into an underestimation of the control reproduction number. Therefore, more careful considerations would be needed for quantitatively robust estimates, but we refrain from expanding on this issue here, given the main purpose of this work is to illustrate the methodology.

Case ascertainment may be an issue more generally. Transmission may not be identifiable if ascertainment rates were to vary over time independently of transmission changes. Moreover, Appendix A.3 shows via simulation that under-reporting of cases, even when constant over duration of reporting, can be an issue, as it appears to dilute the signal of transmission given it leads to smaller case numbers with larger relative intrinsic stochastic noise, and hence rates of changes in the data that are more uncertain. Work will continue to investigate the issues resulting from under-reporting, although again this is likely to be a problem also for the other methods available.

Diseases such as COVID-19 exhibit asymptomatic infection (cases who manifest no symptoms can still infect – or, in case testing is available, cases might infect even prior to being detectable). In this study we have assumed that all infections are detectable for simplicity in illustrating the methodology but in Appendix A.4 we relax this assumption to consider an infectious state prior to potential detection.

Application of `EpiEstim` to real world datasets prior to the COVID-19 pandemic has focussed on shorter time series. Whilst we have not considered the length of the time series in detail, the GAM will require a certain amount of data to ascertain a valid fit and so caution maybe needed in applications to shorter outbreaks than the ones analysed here.

The promising results from this method will be extended to consider simultaneous estimation of parameters across different datasets (i.e. calibrating to death data and case data, or to settings with nested populations, to infer subpopulation-specific reproduction numbers).

This method for estimating the time-varying reproduction number is effective and comparable to other methods. With appropriate handling the method is flexible to the type of epidemic impact measure provided (e.g. mortality or incidence), and it provides an alternative approach to smoothing out surveillance day-of-the-week effects to develop a measure of transmission that can enable action.

## A Supplementary material

### A.1 Approximate Outbreak and Case Estimators

Note that, in the SIS model discussed in Section 2.2.1, when *I*(*t*) ≪ *N* (number of care homes currently experiencing an outbreak small relative to the total number of care homes), then *S*(*t*)*/N* ≈ 1 (i.e. the second term in (7) is approximately 1 and can be ignored) and hence *ρ*(*t*) ≈ *R*_*E*_(*t*), i.e. the control and effective reproduction numbers are approximately the same. In this case, we may derive an approximation 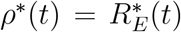 to both. If we additionally assume that enough time has passed so that the contribution of the initial condition *I*(0) can be ignored, (7) further simplifies to

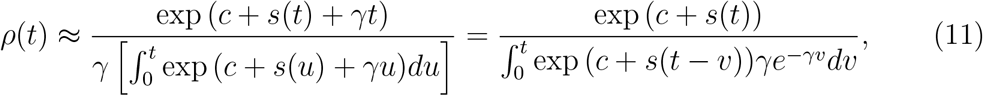

where on the right we have highlighted how the integral at the denominator can be viewed as a convolution of the incidence from the GAM going back into the past (we applied the change of variable *v* = *t* − *u*) with the generation time distribution, which in our model is simply an exponential distribution.

If the generation time distribution had finite support [0, *T* ], the integral would be zero outside of it, i.e. between *T* and *t* (assuming *T < t*). This is not the case for an exponential distribution, but we can still choose *T* large enough that [0, *T* ] contains the the bulk of mass of the generation time distribution. We can then split the integral a more “relevant” part concerning the recent past (*v* ∈ [0, *T* ]) and the rest (*v* ∈ [*T, t*]). With the change of variable *z* = *v* − *T* and using the fact the the incidence exp (*c* + *s*) is bounded by a certain maximum *M*, as it fits observed data, the contribution of the more distant past to the integral is

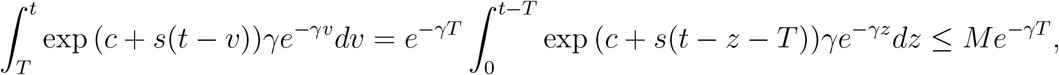

which is small for sufficiently large *T* . Therefore, from (11),

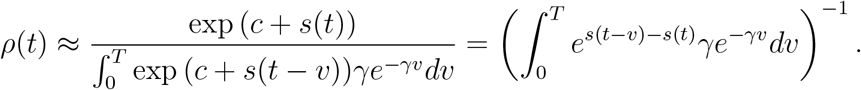

If we Taylor expand 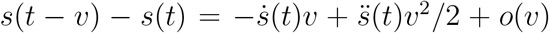 and assume *s* is approximately linear in [0, *T* ] (i.e.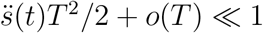), we find

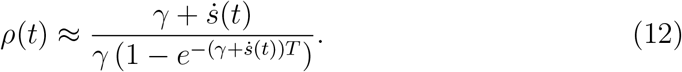

Assuming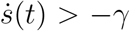, for *T* sufficiently large, the parenthesis at the denominator is approximately 1, and hence

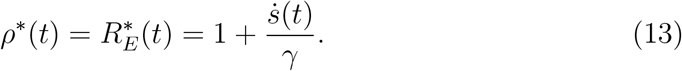

Note that the condition 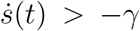 has a natural interpretation, based on the model assumed: if suddenly no transmission occurred (*β*(*t*) = 0), then the currently infected cases would recover at rate *γ* so both prevalence and incidence would decrease exponentially at rate *γ* (i.e. grow at rate −*γ*); therefore, the model would not be able to meaningfully fit a decline in the data with a slope steeper than −*γ*, and in fact if 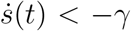 both (12) and (13) yield a negative reproduction number, which is biologically meaningless. This, however, is not a limitation of the estimator *per se*, but rather a problem of model misspecification, i.e. the assumption of an exponentially distributed duration of the infectious period intrinsic in the model formulation would be incompatible with the observed data. In other words, by choosing an SIS Markovian model we are implicitly assuming 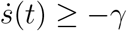. The choice of how large *T* (and similarly *t*) needs to be to control the approximation then depends on how quickly the exponential decay of the generation time distribution makes the incidence going back in the past negligible, which may happen very slowly even for 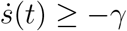 when 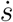 is sufficiently close to −*γ*.

Finally, note that the Taylor expansion used is predicated on the fact *s* should not oscillate too quickly in [*t* − *T, t*]. However, one can strengthen the argument to include cases in which the incidence oscillates quickly around a broadly stable exponential growth at some rate *r*. Then the approximation (13) still applies, but with 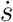 replaced by *r*. However, given *s* is a spline and our goal is to fit the incidence with a relatively “smooth” curve, such cases are less relevant.

An approximation for the estimator used in the SEIRS model of Section 2.2.2 can be obtained following a similar argument to the one described above.

### A.2 Deaths

If only deaths are observed, we can fit the GAM to these data as done in (1) for cases, obtaining

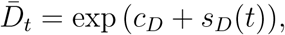

where *c*_*D*_ incorporates (potential) day-of-the-week effects for notation brevity. Substitution into the increment of *D* in the compartment model in Section 2.2.2 gives

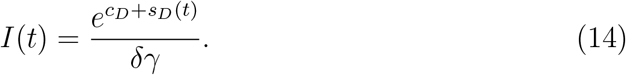

Recalling the definition of *ρ*(*t*) = *β*(*t*)*/ν*, with *ν* from (9) and *β*(*t*) from the equation for *Ė*(*t*) in the ODE system, we obtain Substitution of (14) and its derivative with respect to time into the equation for *İ*(*t*) gives

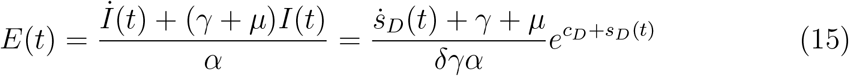

and hence

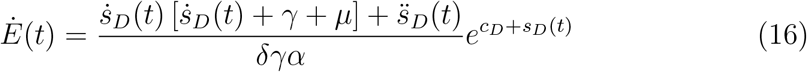

so that

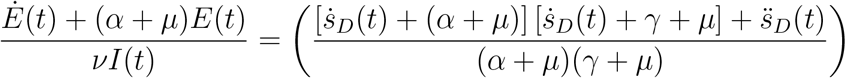

which is also an estimate of the effective reproduction number. Observe that

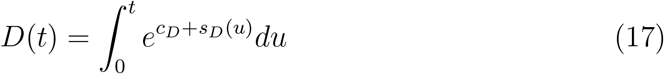

and

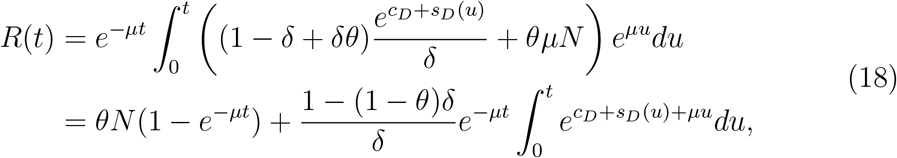

so that finally

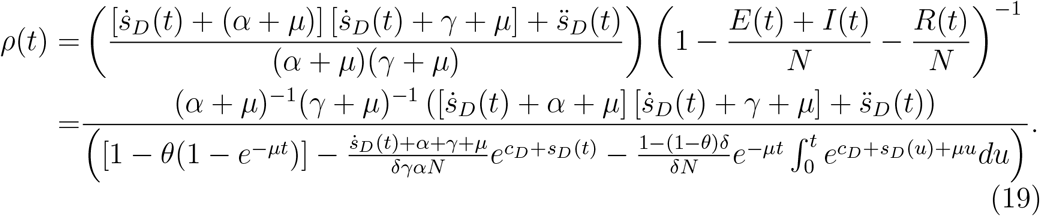

### A.3 Under-reporting in case reporting

The surveillance systems associated with COVID-19 may be expected to almost fully ascertain cases, given testing strategies in place in long-term care facilities from September 2020 when regular whole home testing was implemented. However, not all surveillance systems in place are certain to successfully detect all cases (say due to weak symptoms or mild illness). Hence, data may only reflect a subset of the total epidemic curve.

To investigate under-ascertainment with this framework a binomial sample is used to simulate the daily reported cases with some probability of detection of actual cases. To introduce further noise, a Beta distribution will be used to model the detection probability because the actual detected rate for disease is usually uncertain and likely variable. Therefore we simulate imperfect surveillance using the Beta-Binomial distribution.

Figure 7 shows the reproduction number estimation result of sampling from ‘true’ case numbers (Figure 2 top panel) with different mean detection probabilities. The beta distribution parameters, denoted by *a* and *b*, give a mean *π* = *a/*(*a* + *b*) so that in Figure 7 the range is from *π* = 7*/*9 (pink line) down to 1*/*3 (red line) with the *b* = 2 held constant. The central estimator appears to be weaker as the detection probability drops.

**Figure 7:**
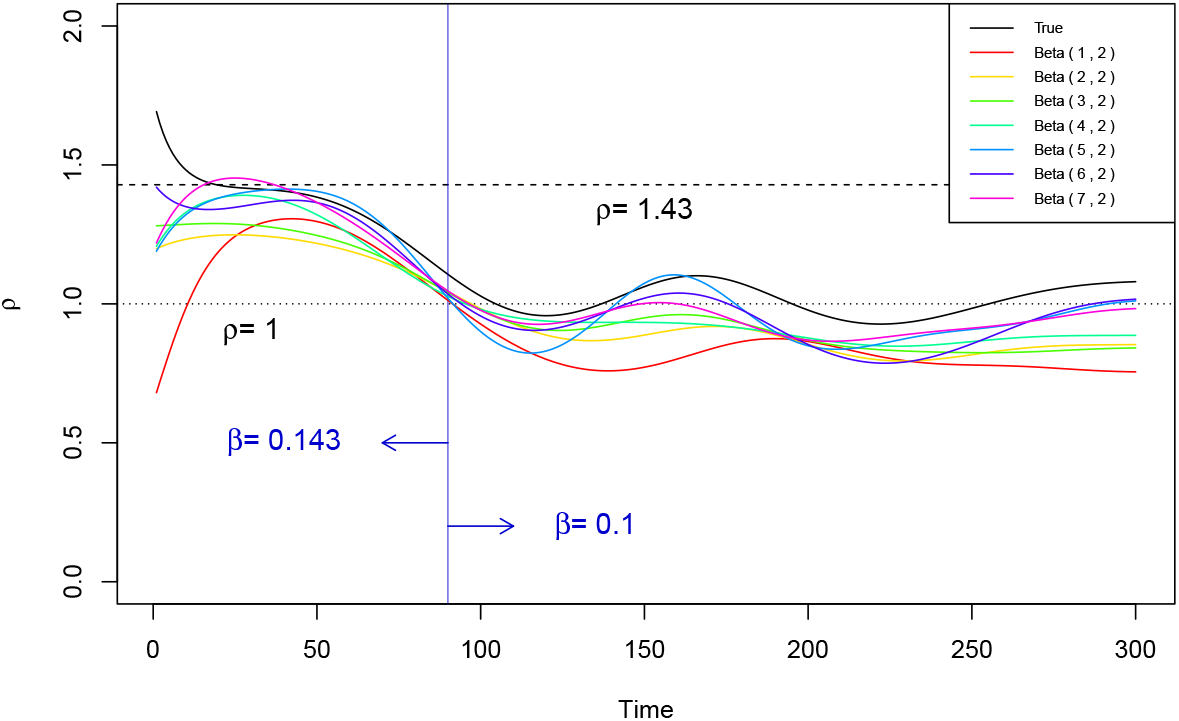
Estimated reproduction numbers *ρ*(*t*) of daily new case from simulated SEIR epidemic (Figure 2 top panel) and seven corresponding Beta-Binomial samples. The black line is estimated from fully-detected daily new cases, while the other lines (coloured lines) are estimated from incompletely detected cases. The predictions of *ρ*(*t*) appear to have been underestimated accordingly (i.e. they are all below the black line, and roughly gradually decrease as the fraction of detected cases decreases).

However, the factors that cause under-reporting are complex so we have little information on the shape of the distribution of detection probability. We may then fix the mean detection to *π* = 1*/*3 but consider different shapes around this mean. For very large values of *b* then the sampled values of *p* are very close to the mean, when *a* = 1 the distribution is triangular (mode at zero) and then for *b <* 1 the distribution becomes ‘U’ shaped with modes at zero and one. In the latter case the probability of detection is very polarised.

An alternative SEIR simulation is shown in Figure 8, but with no transmission-switching intervention in order to ensure a clearer visualisation of the impact of under-reporting. The crosses represent the simulated observations of daily new cases and the curves are GAM fits. In Figure 9, the reproduction number and its credible interval for the full data (black line) provides a reasonably accurate estimate only when simulated case numbers are above 20 (day 50-150). The simulated sampling with mean of 1/3 is good enough only when the Beta parameters are greater than 1, and even in these cases estimates are reliable only up to peak, tending to lower values than those expected afterwards. We note however, that the early phase of the outbreak is most likely the period of greatest interest. A very polarised ascertainment (i.e. red lines) suggests the poorest estimate. In conclusion, these simulations suggest that low case ascertainment may lead to biased estimates. We deemed this somewhat surprising, as under-reported case incidence is generally not expected to result in underestimation of the reproduction number [12], although this expectation might likely be a consequence of assumed stable ascertainment and relatively high incidence. The simulations above, instead, are stochastic and based on fairly small populations (10,000 people in total) and therefore produce low-count cases as well as ‘exaggerated’ missingness to deliberately stress the method. The GAM fitting has imposed a smoothing process on this low case incidence data for a given noise model, so the loss of information (or rather than signal-to-noise ratio) requires further investigation.

**Figure 8:**
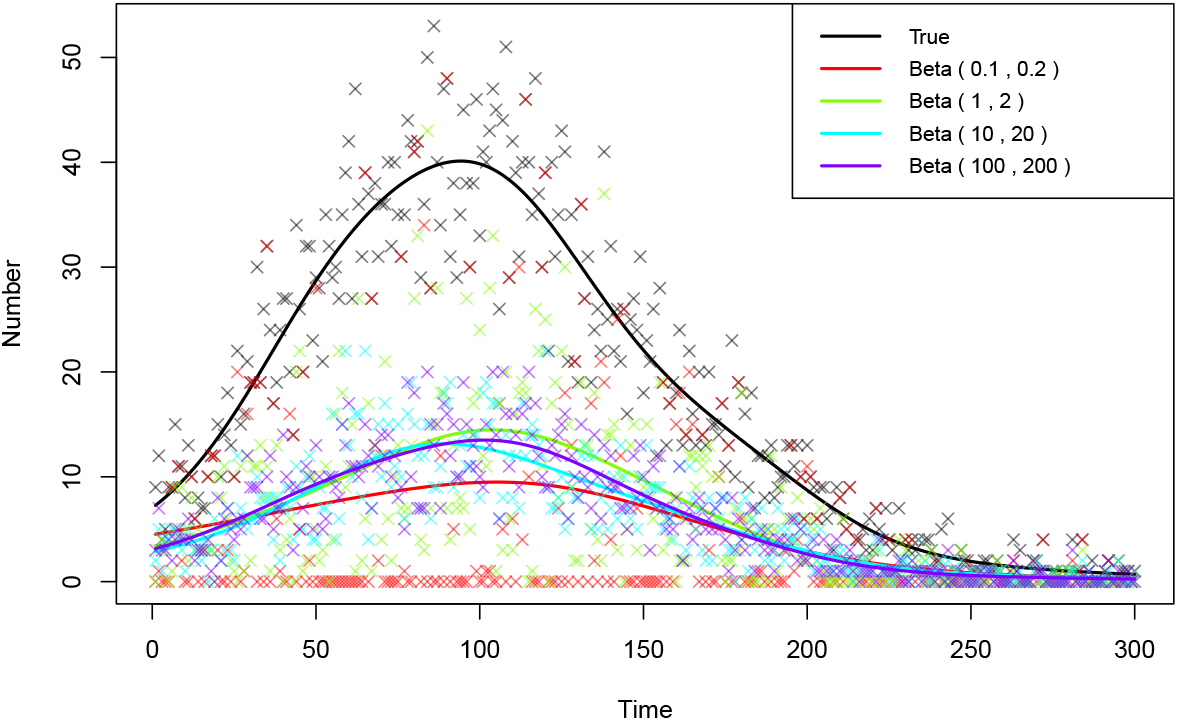
GAM fits, with option `family` = `quasipoisson`, on simulated case data (black) and four Beta-Binomial samples with detected probability mean *π* = 1*/*3 (red: Beta(0.1, 0.2); green: Beta(1, 2); cyan: Beta(10, 20); and purple: Beta(100, 200)). The daily new case data are generated from the simulated stochastic SEIR model without intervention, with parameters *N* = 10^4^, *E*(0) = *I*(0) = 50, *α* = 1*/*7, *γ* = 0.1, Δ*t* = 0.15.

**Figure 9:**
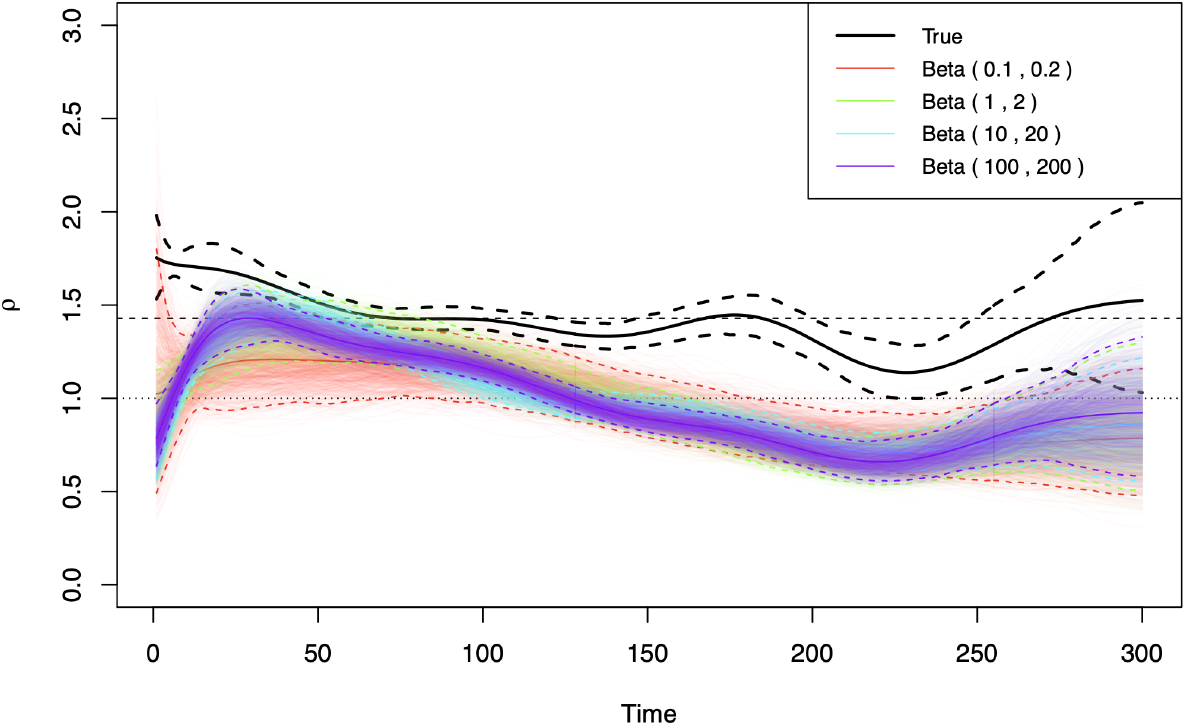
95% Credible Interval for the reproduction number estimated from the data generated with a Beta-distributed detection probability with mean *π* = 1*/*3 (Figure 8). The black solid line is estimated from the fully-detected daily new cases (black GAM fit in Figure 8), with black dashed lines denoting the CrI, while the coloured lines are estimated from incompletely-detected cases (red lines: Beta(0.1, 0.2); green: Beta(1, 2); cyan: Beta(10, 20); and purple: Beta(100, 200)).

### A.4 A model with pre-symptomatic infectivity

In the COVID-19 pandemic, many cases started being infectious before showing symptoms. To capture this, an additional state of infection can be added (*P* for pre-symptomatic, or prodromal) to the disease spread model, leading to the system

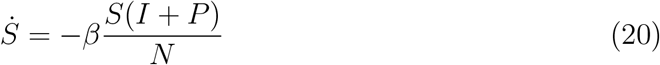

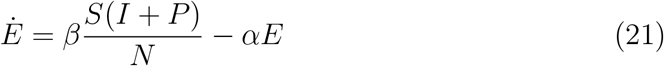

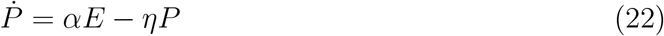

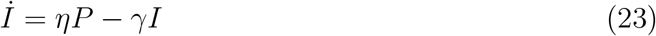

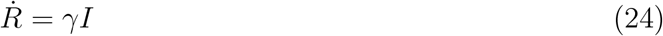

This model assumes that *S* are those susceptible, *E* are those infected but not infectious, *P* are those infected and infectious but not yet detectable, *I* are those infected, infectious and detectable and *R* are those recovered and immune. Crucially it assumes that the disease states *P* and *I* are equally infectious. Here, we have phrased the motivation in terms of symptoms but the logic remains for situations of widespread availability of rapid diagnostic testing and diseases (such as COVID-19) where cases are infectious prior to detectability by test: in this case, then, the interpretation can be changed to *P* being undetectable infectious cases and *I* detectable infectious ones.

The new disease incidence is given by those entering *I* state, i.e. *ηP* in (23), and we may write disease states in terms of the GAM estimators *c* and *s*(*t*):

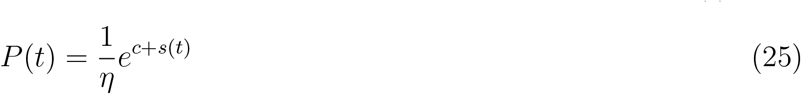

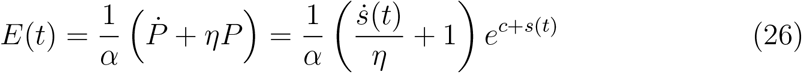

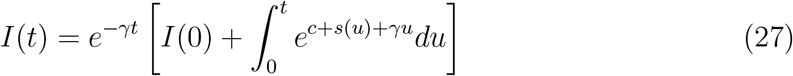

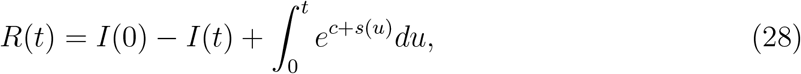

where the last equation was derived by replacing *γI*(*t*) with 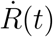 in the equation for *İ*(*t*) above and solving for *R*(*t*) and assuming for simplicity *R*(0) = 0. The control reproduction number for such a system (e.g. compare with the basic reproduction number in [43]) is

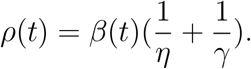

Defining the timescale parameter 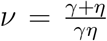 so that, as usual, *ρ*(*t*) = *β*(*t*)*/ν*, and expressing *β*(*t*) from the equation for *Ė*(*t*) above, we obtain

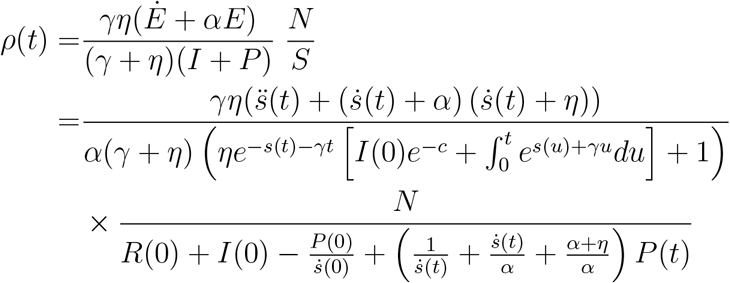

and dropping the *N/S* factor we obtain the effective reproduction number. This proof-of-principle shows that it is possible to derive estimators for more complex transmission model structures, although we do not expand in this direction here but rather leave this derivation as a basis for further analysis and development of the tools.

## Data Availability

All data produced in the present study are available upon reasonable request to the authors

## B Acknowledgements

We would like to thank Chris Overton for useful discussions about serial intervals. LP gratefully acknowledges the Wellcome Trust and Royal Society (grant 202562/Z/16/Z). LP, TH and IH are also supported by the JUNIPER modelling consortium (grant MR/V038613/1), by the Alan Turing Institute for Data Science and Artificial Intelligence under the EPSRC grants EP/N510129/1 and EP/V027468/1 and by the UKRI Impact Acceleration Account (IAA 386). YH, LP, TH and IH also acknowledge the UK Health Security Agency (UKHSA) for honorary contracts and funding. The views expressed are those of the author(s) and not necessarily those of the Department of Health or UKHSA.

